# Kindergarten MMR Coverage, Vaccination Exemptions, and State-Level Measles Incidence in the United States, 2024–2026

**DOI:** 10.64898/2026.11.17.26368302

**Authors:** Sumon Ghosh, Thankam S. Sunil, Sukanta Chowdhury, Mohammad Nayeem Hasan, Md Khademul Islam, Job K. Ronoh, Joseph M. Kungu, Dennis N. Makau

**Affiliations:** Department of Biomedical and Diagnostic Sciences, University of Tennessee, Knoxville, TN 37996, USA; Department of Nutrition and Public Health Sciences, University of Tennessee, Knoxville, TN 37996, USA; International Centre for Diarrhoeal Disease Research, Bangladesh (icddr,b), Dhaka 1212, Bangladesh; Monitoring, Evaluation & Research Department, Community Partners International, East Dolphin Point, Cox’s Bazar 4700, Bangladesh; College of Veterinary Medicine, Animal Resources and Biosecurity, Makerere University, Kampala, Uganda; Department of Public Health Pharmacology and Toxicology, Faculty of Veterinary Sciences, University of Nairobi, P. O. Box 29053-00625 Kangemi, Kenya

**Keywords:** Measles, measles-mumps-rubella vaccination, vaccination exemptions, kindergarten vaccination coverage, measles incidence, United States

## Abstract

**Objectives:** To examine associations of kindergarten measles-mumps-rubella (MMR) coverage and vaccination exemptions with state-level measles incidence in the United States, 2024–2026.

**Methods:** In this ecological study of 50 states and Washington, DC (up to 153 jurisdiction-years), we linked CDC surveillance, SchoolVaxView, and Census data. Negative-binomial models estimated incidence rate ratios (IRRs) per percentage-point increase for completed years 2024–2025 (generalized estimating equations) and through MMWR week 39, 2024–2026 (maximum likelihood, adjusted for density and poverty).

**Results:** Cases rose from 269 (2024) to 2,263 (2025); 3,642 were reported provisionally through October 2, 2026. Completed-year IRRs were 0.875 (95% CI=0.806, 0.950) for MMR coverage, 1.202 (1.075, 1.344) for any exemption, and 1.207 (1.059, 1.376) for nonmedical exemption. Adjusted week-39 IRRs were 0.945 (0.837, 1.068), 1.147 (1.004, 1.310), and 1.131 (0.972, 1.316). Under cluster-robust maximum likelihood, completed-year intervals for MMR coverage and nonmedical exemption included 1.

**Conclusions:** Higher exemption prevalence was associated with greater measles incidence; other estimates were imprecise.

**Public Health Implications:** Monitoring exemptions alongside MMR coverage may help identify jurisdictions needing closer vaccination-gap assessment. Ecological findings do not establish causality.

## Introduction

Measles is one of the most contagious human infectious diseases and remains a major vaccine-preventable disease worldwide^1,2^. Sustained high population immunity can limit onward transmission following measles importation, whereas spatially clustered pockets of susceptible individuals can permit sustained transmission and large outbreaks even when vaccination coverage appears high at the state or national level. Two-dose measles-containing vaccine coverage of approximately 95% is generally considered an important population-level benchmark for preventing sustained transmission; however, aggregate coverage estimates can mask substantial local heterogeneity in vaccination and susceptibility^3-5^.

Concurrently, kindergarten vaccination surveillance has documented a sustained decline in U.S. MMR coverage. Nationally, two-dose MMR coverage among kindergartners decreased from 95.2% in the 2019–2020 school year to 92.7% in 2023–2024 and 92.4% in 2025–2026. During the same period, exemptions from one or more required vaccines increased, reaching 4.2% nationally in 2025– 2026^6,7^. These domestic trends are occurring amid a broader global resurgence of measles. WHO and CDC estimated that 10.3 million measles cases occurred worldwide in 2023, when global coverage with the first and second doses of measles-containing vaccine was 83% and 74%, respectively^8^. In the United States, recent state-level analyses have demonstrated a strong inverse association between nonmedical exemption prevalence and kindergarten MMR vaccination coverage^9-11^. Previous studies have also linked nonmedical exemptions to increased individual and community risk of measles, while state exemption policies have been associated with differences in kindergarten vaccination coverage^12,13^.

Within the U.S., recent measles burden has been highly geographically concentrated, with a relatively small number of states accounting for a disproportionate share of reported cases. Between January 1 and April 17, 2025, the CDC reported 800 confirmed measles cases in the United States, including 654 (82%) associated with a multistate outbreak in New Mexico, Oklahoma, and Texas. Most cases (96%) occurred among unvaccinated individuals or those with unknown vaccination status^14,15^. This geographic concentration underscores an important analytic challenge: crude state-level case counts reflect not only underlying population susceptibility but also differences in population size, international importation, and the occurrence and localized outbreak dynamics.

We therefore aimed to (1) describe state-level measles burden during 2024–2026 using counts and population-standardized incidence; (2) evaluate associations of kindergarten MMR coverage and vaccination exemptions with state-level measles rates during completed calendar years; and (3) examine the robustness of these associations when incorporating 2026 data through MMWR week 39 and accounting for contextual factors, outbreak concentration, and surveillance-related assumptions.

## Methods

### Study Design and Data Sources

We conducted a state-level ecological study of measles incidence and kindergarten vaccination indicators across 51 U.S. jurisdictions (50 U.S. states and the District of Columbia) during 2024– 2026. The analytical datasets were structured in long format, with one observation per jurisdiction-year. The completed-year surveillance dataset comprised 102 jurisdiction-year observations for 2024–2025 (51 jurisdictions × 2 years), whereas the comparable-period NNDSS dataset comprised 153 jurisdiction-year observations for 2024–2026 (51 jurisdictions × 3 years). All 51 jurisdictions were represented in each applicable year in both surveillance datasets, yielding balanced source panels with consistent geographic coverage. New York City surveillance records were combined with New York State to maintain consistent geographic units.

Two distinct CDC measles surveillance data sources were analyzed separately: (1) reconciled measles case-history and state-level surveillance files for completed calendar years 2024–2025, downloaded on October 2, 2026; and (2) provisional National Notifiable Diseases Surveillance System (NNDSS) Weekly Data for 2024–2026, downloaded on October 8, 2026. The completed-year surveillance files were used to estimate annual measles incidence for 2024 and 2025. To incorporate 2026 under comparable surveillance follow-up, we extracted cumulative counts of total, indigenous, and imported measles from the NNDSS Weekly Data through Morbidity and Mortality Weekly Report (MMWR) week 39 in each year from 2024 through 2026. MMWR week 38 was examined as a surveillance-cutoff sensitivity analysis. The reconciled completed-year CDC surveillance files and the provisional NNDSS Weekly Data were analyzed separately and were not combined because they represent different surveillance periods and are subject to different reporting and revision processes^16^.

In addition, kindergarten measles-mumps-rubella (MMR) vaccination coverage and exemption prevalence data were obtained from CDC SchoolVaxView. State population estimates, 2020 population density, and poverty prevalence from the 2023 and 2024 American Community Survey (ACS) were obtained from the US Census Bureau. All analyses used publicly available, aggregated data.

### Outcomes and Vaccination Indicators

The primary outcome was the number of reported measles cases within each jurisdiction-year. For the NNDSS analyses, cases were classified as imported or indigenous according to CDC reporting categories; total cases represented the sum of both. State-level measles incidence was expressed per million population using corresponding population denominators.

The principal exposures were kindergarten MMR vaccination coverage, any vaccine exemption, and nonmedical exemption, each expressed as a percentage of kindergartners. Any exemption represented exemption from at least one school-required vaccine and was not necessarily specific to MMR^6,7^. Vaccination indicators from the school year ending in each calendar year were linked to the corresponding calendar-year measles outcomes to maintain consistent temporal alignment across jurisdictions. Because these measures were not necessarily recorded before all reported measles cases, the associations were interpreted as contemporaneous rather than prospective. Preceding-school-year vaccination indicators were evaluated in sensitivity analyses to assess the robustness of findings to exposure timing. Each vaccination indicator was evaluated in a separate model. Missing vaccination indicator values were retained as missing and were not recoded as zero or imputed. Jurisdiction-year observations with unavailable values for a given vaccination indicator were excluded only from the corresponding exposure-specific regression model.

### Statistical Analysis

We summarized annual and provisional measles counts, state-level incidence, and geographic variation. Associations between kindergarten vaccination indicators and measles incidence were estimated using negative-binomial regression models to accommodate potential overdispersion in measles case counts arising from substantial heterogeneity in outbreak burden across jurisdictions. Models included a logarithmic link, the logarithm of the jurisdiction population as an offset, and calendar-year indicators. Results were expressed as incidence rate ratios (IRRs) and 95% confidence intervals (CIs) per 1-percentage-point increase in each vaccination indicator.

The primary analysis examined associations between kindergarten vaccination indicators and state-level measles incidence using completed-year surveillance data for 2024–2025. A complementary analysis used surveillance counts through MMWR week 39 for each year from 2024 through 2026 to evaluate associations across comparable reporting periods while incorporating available 2026 data. The two datasets were analyzed separately because they represent different surveillance windows. Although both source surveillance panels were balanced, the exposure-specific analytical panels were unbalanced because of missing vaccination indicators. The completed-year models included 99 jurisdiction-year observations from 50 jurisdictions for MMR coverage and any vaccination exemption, and 93 observations from 47 jurisdictions for nonmedical exemption. The corresponding comparable-period week-39 models included 148, 149, and 139 jurisdiction-year observations from 50, 50, and 47 jurisdictions, respectively.

For completed calendar years 2024–2025, we fitted negative-binomial generalized estimating equations (GEE) with an exchangeable working correlation structure and jurisdiction-clustered robust sandwich standard errors to estimate population-averaged associations. The negative-binomial dispersion parameter was estimated separately using maximum-likelihood regression and held fixed during GEE estimation. Models included calendar-year indicators and a log population offset. A negative-binomial NB2 maximum-likelihood model with jurisdiction-clustered robust standard errors was evaluated as an estimator-sensitivity analysis. For the comparable-period NNDSS analysis covering 2024–2026 through MMWR week 39, we used negative-binomial NB2 maximum-likelihood regression with jurisdiction-clustered robust standard errors. The two analyses were fitted separately using their respective CDC surveillance datasets. The completed-year GEE analysis was used to summarize population-averaged associations while accounting for repeated jurisdiction-level observations; the comparable-period NB2 maximum-likelihood analysis was retained as a complementary specification for the separately extracted provisional surveillance series.

To assess potential confounding by geographic and socioeconomic characteristics, comparable-period models were additionally adjusted for log-transformed 2020 state population density and ACS poverty prevalence. The primary poverty specification assigned 2023 ACS estimates to 2024 outcomes and 2024 ACS estimates to 2025–2026 outcomes. Calendar-year-adjusted and fully contextually adjusted models were compared within the same exposure-specific analytic samples. Measles case counts and population denominators were complete across both source panels, and population density and poverty prevalence were complete in the comparable-period analytical dataset. Thus, exclusions from the principal regression models were attributable to unavailable vaccination indicators rather than missing outcome or contextual covariate data.

Sensitivity analyses examined week-38 surveillance counts, preceding-school-year vaccination indicators, alternative poverty-year assignments, and statistical estimation approaches. For the completed-year analysis, negative-binomial GEE estimates were compared with NB2 maximum-likelihood estimates using jurisdiction-clustered robust standard errors to assess sensitivity to the estimation approach. Completed-year analyses additionally assessed the exclusion of four high-burden states and restriction to jurisdiction-years with at least 90% SchoolVaxView survey completeness. Model convergence and dispersion were evaluated. All statistical tests were two-sided; estimates were interpreted as ecological associations rather than causal effects. All analyses were performed using R statistical software (version 4.6.0; R Foundation for Statistical Computing, Vienna, Austria).

## Results

### Descriptive epidemiology

Measles activity increased markedly during the study period, with reconciled case counts rising from 269 cases in 2024 to 2,263 in 2025. An additional 3,642 cases were reported in the provisional year-to-date (YTD) dataset available on the October 2, 2026 analytic cutoff. Because the 2026 observation period was incomplete, its cumulative count was interpreted descriptively rather than as a completed annual total. The number of states and the District of Columbia reporting at least one case increased from 31 in 2024 to 44 in 2025 and 46 in 2026 YTD, with the median state-level incidence increasing from 0.160 per million population in 2024 to 1.396 in 2025 and 1.971 in 2026 YTD (Table S1).

The geographic distribution of reported cases and population-adjusted incidence varied substantially across years (Figure 1; Figure S1). South Carolina, Texas, Pennsylvania, and Utah together accounted for 58.24% of cumulative cases, highlighting the concentration of measles burden within a small number of jurisdictions. Consistent with this pattern, the Gini coefficient for cumulative cases across states and the District of Columbia was 0.741 (Figure S2); State-specific counts for the 15 highest-burden states are presented in Supplementary Table S2.

**Figure 1.**
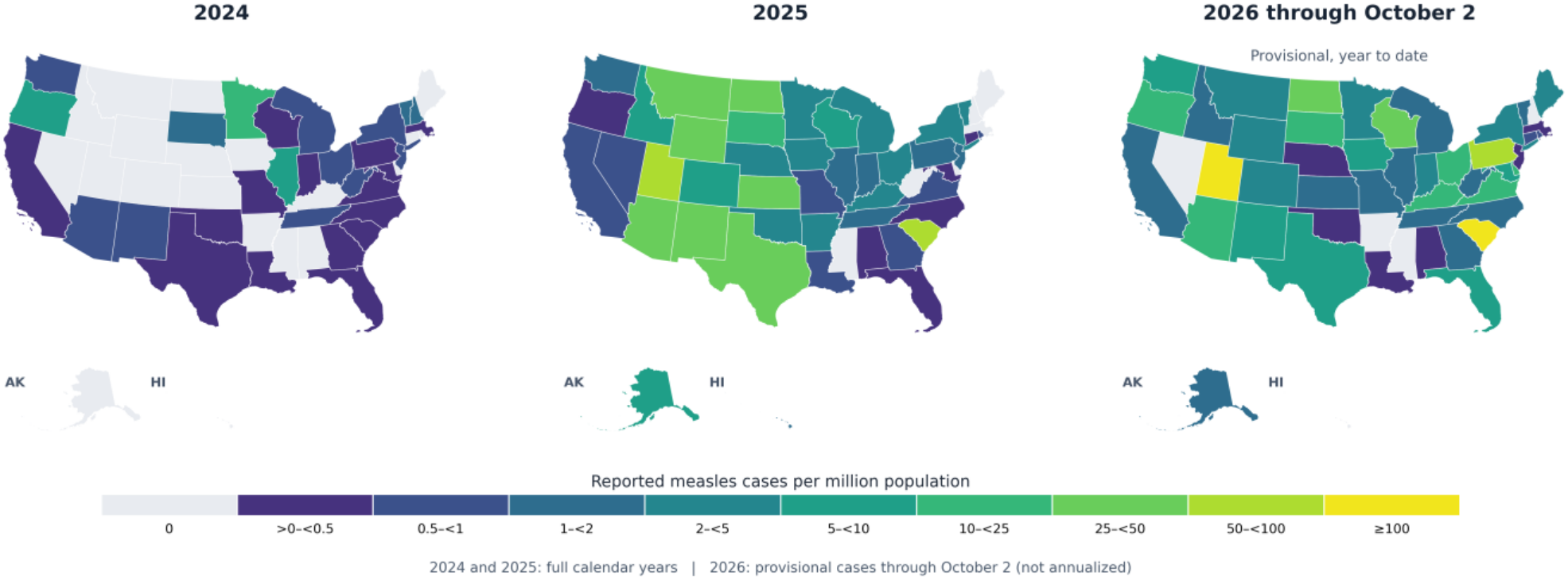
State-Level Reported Measles Incidence per Million Population, United States, 2024–2026. Maps show reported measles incidence during completed calendar years 2024 and 2025 and provisional cumulative incidence through October 2, 2026. The 2026 estimates are not annualized. New York includes New York City. Alaska and Hawaii are repositioned for display. State boundaries were obtained from the 2025 US Census Bureau TIGER/Line shapefiles. Maps use the October 2 surveillance series, distinct from the NNDSS comparable-period regression dataset.

### Vaccination coverage and exemption patterns

The 2025-2026 SchoolVaxView data showed substantial between-state variation in kindergarten MMR coverage, any vaccination exemption prevalence, and nonmedical exemption prevalence (Figure S3). Because exemption estimates refer to exemption from one or more required vaccines and are not necessarily MMR-specific, the geographic patterns in Figure S3 should be interpreted as indicators of the broader school-vaccination environment rather than as direct measures of MMR-specific exemption.

### Primary completed-year regression analysis

In the primary 2024–2025 negative-binomial GEE models accounting for repeated observations within jurisdictions, higher kindergarten MMR coverage was associated with a lower state-level measles rate. Each 1-percentage-point increase in MMR coverage was associated with a 12.5% lower rate (IRR 0.875, 95% CI 0.806-0.950; p=0.001). In separate models, each 1-percentage-point increase in any vaccination exemption was associated with a 20.2% higher measles rate (IRR 1.202, 95% CI 1.075-1.344; p=0.001), and each 1-percentage-point increase in nonmedical exemption prevalence was associated with a 20.7% higher rate (IRR 1.207, 95% CI 1.059-1.376; p=0.005) (Table 1). These estimates should be interpreted separately because the vaccination and exemption measures were highly correlated.

**Table 1.** Associations of Kindergarten Vaccination Indicators With Measles Incidence From Negative-Binomial Generalized Estimating Equation Models: United States, 2024–2025.

| Exposure (per 1 percentage point) | Jurisdiction-year N | IRR | 95% CI | p-value |
|---|---|---|---|---|
| MMR coverage | 99 | 0.875 | 0.806-0.950 | 0.001 |
| Any exemption | 99 | 1.202 | 1.075-1.344 | 0.001 |
| Nonmedical exemption | 93 | 1.207 | 1.059-1.376 | 0.005 |

Note. IRR = incidence rate ratio; CI = confidence interval; MMR = measles-mumps-rubella. Estimates represent associations per 1-percentage-point increase in each vaccination indicator. Separate negative-binomial GEE models included calendar-year indicators and a log population offset, with jurisdiction-level clustering, an exchangeable working correlation structure, and robust sandwich standard errors. Negative-binomial dispersion was estimated separately and held fixed during GEE estimation. Sample sizes reflect available exposure data.

Estimator-sensitivity analyses yielded similar point estimates but wider confidence intervals under NB2 maximum likelihood; intervals for MMR coverage and nonmedical exemption included 1 (Table S3).

### Comparable-Period and Robustness Analyses

Sensitivity analyses using preceding-school-year vaccination indicators yielded associations consistent in direction with the primary completed-year models (Table S4). Excluding the four highest-burden states (Texas, South Carolina, Arizona, and Utah) attenuated the associations, although their directions remained consistent (Table S5). Restricting analyses to jurisdiction-years with at least 90% SchoolVaxView survey completeness produced similar estimates (Table S6).

In the comparable-period analysis using NNDSS surveillance data through MMWR week 39 in 2024–2026, higher kindergarten exemption prevalence was associated with greater total measles incidence. Each 1-percentage-point increase in any exemption was associated with an 18.1% higher measles rate (IRR = 1.181; 95% CI = 1.045, 1.336), while nonmedical exemption was associated with an 18.0% higher rate (IRR = 1.180; 95% CI = 1.018, 1.369). Higher MMR coverage was associated with lower measles incidence, although the confidence interval included 1 (IRR = 0.903; 95% CI = 0.794, 1.029). For indigenous measles, any exemption (IRR = 1.210; 95% CI = 1.058, 1.384) and nonmedical exemption (IRR = 1.198; 95% CI = 1.025, 1.399) were positively associated with incidence. For imported measles, effect estimates were closer to the null, and confidence intervals included 1 for all three vaccination indicators (Table 2; Table S7 and S8).

**Table 2.** Associations of kindergarten vaccination indicators with State-Level measles incidence through MMWR week 39, United States, 2024–2026.

| Outcome / exposure | Calendar-year adjusted IRR (95% CI) | Contextually adjusted IRR (95% CI) | Adjusted p-value |
|---|---|---|---|
| **Total measles** |  |  |  |
| MMR coverage | 0.903 (0.794-1.029) | 0.945 (0.837-1.068) | 0.365 |
| Any exemption | 1.181 (1.045-1.336) | 1.147 (1.004-1.310) | 0.044 |
| Nonmedical exemption | 1.180 (1.018-1.369) | 1.131 (0.972-1.316) | 0.112 |
| **Indigenous measles** |  |  |  |
| MMR coverage | 0.881 (0.759-1.023) | 0.940 (0.818-1.080) | 0.379 |
| Any exemption | 1.210 (1.058-1.384) | 1.145 (0.989-1.327) | 0.071 |
| Nonmedical exemption | 1.198 (1.025-1.399) | 1.116 (0.951-1.311) | 0.179 |
| **Imported measles** |  |  |  |
| MMR coverage | 0.984 (0.908-1.067) | 1.013 (0.936-1.096) | 0.746 |
| Any exemption | 1.011 (0.923-1.109) | 0.967 (0.874-1.071) | 0.521 |
| Nonmedical exemption | 1.057 (0.952-1.174) | 1.019 (0.904-1.150) | 0.756 |

Note. IRRs represent the association with a 1-percentage-point increase in each vaccination indicator. Calendar-year-adjusted models include calendar-year indicators and a log population offset. Census-adjusted models additionally include log-transformed 2020 Census population density and preceding-available American Community Survey poverty prevalence. State-clustered robust standard errors were used. MMR coverage models included 148 Jurisdiction-year observations; any-exemption models, 149; and nonmedical-exemption models, 139.

Adjustment for state-level population density and poverty prevalence attenuated most associations with total measles incidence. Any vaccination exemption remained positively associated with incidence (adjusted IRR = 1.147; 95% CI = 1.004, 1.310), whereas confidence intervals for MMR coverage and nonmedical exemption included 1. Associations with indigenous measles were also attenuated, with adjusted confidence intervals including 1 for all three vaccination indicators. Imported-measles estimates remained near the null (Table 2; Table S9; Figures 2 and S4).

**Figure 2.**
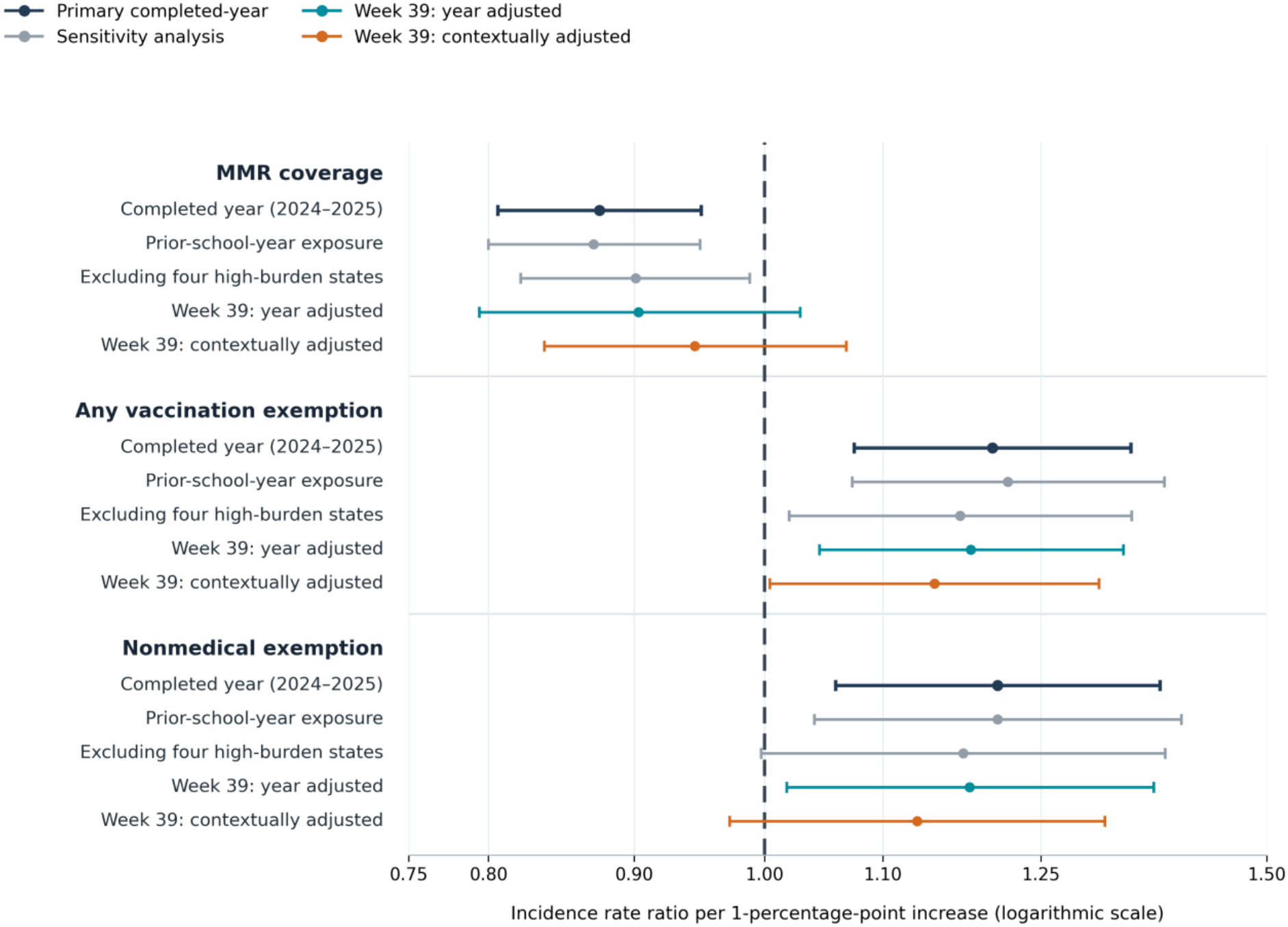
Associations of Kindergarten Vaccination Indicators With Measles Incidence Across Analytical Specifications: United States, 2024–2026. **Note**. Points represent incidence rate ratios (IRRs), and horizontal lines represent 95% confidence intervals per 1-percentage-point increase in each vaccination indicator. The vertical dashed line indicates the null value (IRR = 1.0). Completed-year estimates were obtained from CDC surveillance data for 2024–2025; comparable-period estimates were obtained from provisional National Notifiable Diseases Surveillance System data through Morbidity and Mortality Weekly Report week 39 for 2024–2026. Surveillance datasets were analyzed separately. Completed-year primary estimates were obtained using negative-binomial GEE, whereas comparable-period estimates were obtained using NB2 maximum-likelihood regression with jurisdiction-clustered robust standard errors. Contextually adjusted models additionally included population density and poverty prevalence. Sensitivity analyses evaluated preceding-school-year vaccination indicators and exclusion of four high-burden states. The horizontal axis uses a logarithmic scale.

### Sensitivity to Poverty Measurement and Exposure Timing

Alternative specifications of poverty prevalence had little effect on the findings. Using fixed 2023 poverty estimates, the adjusted association between any exemption and total measles incidence remained positive (IRR = 1.163; 95% CI = 1.021, 1.324), compared with 1.147 (95% CI = 1.004, 1.310) in the primary contextual-adjustment specification (Table S10). Analyses using preceding school year vaccination and exemption indicators yielded similar patterns of association with total and indigenous measles incidence, although these models did not include Census-derived contextual covariates (Table S11).

## Discussion

This ecological analysis identified increasing reported measles burden during 2024–2026 and associations between kindergarten vaccination indicators and state-level incidence. Higher MMR coverage was associated with lower incidence in the completed-year analysis, whereas higher exemption prevalence was associated with greater incidence. In the comparable-period analysis, contextual adjustment attenuated most associations; only any vaccination exemption retained a confidence interval excluding the null. These findings suggest that exemption prevalence may help identify jurisdictions requiring closer assessment of vaccination gaps, although the ecological associations do not establish causality.

The inverse association between kindergarten MMR coverage and measles incidence in the completed-year analysis is consistent with the established importance of high population immunity in limiting measles transmission^17^. CDC identifies approximately 95% two-dose measles vaccination coverage as an important community-level benchmark, although aggregate coverage can conceal localized susceptibility^18^. Previous studies have demonstrated that geographic clustering of unvaccinated individuals increases measles outbreak potential, even where broader coverage estimates appear adequate. Recent outbreaks in Texas, New Mexico, and Oklahoma provide further context, with transmission concentrated in communities with low vaccination coverage and most reported patients being unvaccinated or having unknown vaccination status^14,19^. However, kindergarten MMR coverage reflects vaccination status in a single school-entry cohort rather than population immunity, and state-level averages may mask pockets of susceptibility. These limitations may partly explain the weaker association observed in the comparable-period analysis. Because the completed-year and comparable-period analyses used different surveillance datasets and analytic approaches, differences between their estimates should not be overinterpreted.

Vaccination exemption prevalence showed generally positive associations with measles incidence across analytic specifications. Estimates for any exemption remained positive in analyses using preceding-school-year exposures, excluding the four highest-burden states, restricting to jurisdictions with high survey completeness, and using an alternative NNDSS surveillance cutoff. Nonmedical exemption estimates were similar in direction, although their precision varied and some confidence intervals included the null. These findings are consistent with earlier research linking vaccination exemptions and their geographic concentration to increased measles risk^12,20,21^. Studies of school immunization policies have also associated permissive exemption provisions with increased exemption prevalence and vaccine-preventable disease incidence, whereas repeal of nonmedical exemptions has been associated with improved kindergarten vaccination coverage^9,22^. Our analysis extends this literature by examining school-entry indicators alongside reported measles incidence during renewed US transmission. Nevertheless, because vaccination coverage and exemption measures are correlated and were evaluated in separate models, their estimates should be interpreted as alternative indicators of the school-vaccination environment rather than independent or additive effects.

Variation across surveillance periods, high-burden-state exclusions, and contextual adjustment highlights the sensitivity of ecological associations to outbreak distribution and model specification. Before contextual adjustment, exemption prevalence was positively associated with indigenous measles incidence, whereas imported-measles estimates were closer to the null. After adjustment for population density and poverty prevalence, the estimated associations between exemption prevalence and indigenous measles incidence remained positive but were closer to the null, with confidence intervals that included 1. This pattern raises the possibility that local vaccination conditions may be more closely associated with transmission following importation than with importation itself.

However, the indigenous surveillance category includes cases with unknown location of acquisition. The study was not designed to test whether associations differed significantly between indigenous and imported measles. Finer-scale investigations incorporating vaccination patterns, outbreak linkages, and importation histories are needed to examine this possibility.

Geographic analyses showed substantial variation in measles burden across jurisdictions (Figure S1), with cumulative case counts highly concentrated (Gini coefficient = 0.741; Figure S2). However, this coefficient describes inequality in absolute case counts rather than population-adjusted incidence or transmission risk. Because cumulative counts combine completed-year and provisional surveillance data, state rankings should not be interpreted as stable measures of risk. These findings support finer-scale assessments of vaccination gaps and outbreak settings^4,21^.

Completed-year GEE and NB2 maximum-likelihood models yielded similar point estimates but different confidence intervals. Under NB2 maximum likelihood, intervals for MMR coverage and nonmedical exemption included the null, emphasizing sensitivity to estimation methods and uncertainty in statistical precision (Table S3).

### Strengths and Limitations

This study integrated measles surveillance, kindergarten vaccination indicators, and population data across the 50 states and District of Columbia. Separate completed-year and comparable-period analyses incorporated provisional 2026 surveillance without treating partial-year observations as completed annual counts. Sensitivity analyses evaluated exposure timing, influential states, survey completeness, surveillance cutoffs, statistical estimators, and contextual adjustment.

Several limitations warrant consideration. First, the ecological design precludes individual-level inference or causal interpretation. Kindergarten vaccination indicators do not measure immunity across age groups or within outbreak-affected communities. In addition, the SchoolVaxView any-exemption measure includes exemptions from one or more required vaccines and is not necessarily MMR-specific. Second, differences in school-entry requirements, data collection, reporting practices, and survey completeness may affect comparability across jurisdictions^6,7,23^. Additionally, contemporaneous school-year vaccination indicators may not precede all measles cases within the corresponding calendar year, limiting temporal interpretation. Third, the limited study period, geographic concentration of outbreaks, and provisional surveillance reporting may affect estimate stability and introduce measurement uncertainty. Fourth, contextual adjustment for population density and poverty prevalence was limited to the comparable-period analyses and could not eliminate residual confounding from importation pressure, population mobility, localized vaccination patterns, outbreak settings, and public health responses. Population density was measured using 2020 data, and the available poverty estimates did not provide a distinct preceding-year measurement for 2026. The study was not preregistered; the primary and complementary analysis designations were not prespecified in a registered protocol. Confidence interval precision varied across analytical specifications, and some imported-measles models approached the Poisson dispersion boundary. Inference for the completed-year associations was sensitive to the estimation approach, with wider confidence intervals under the NB2 maximum-likelihood specification than under GEE. Accordingly, the findings should be interpreted as exploratory population-level associations rather than evidence of individual-level or causal effects.

### Public Health Implications

#### Practice

State and local health departments could review school-entry exemption prevalence alongside MMR coverage when identifying jurisdictions for closer assessment of vaccination gaps, outbreak readiness, and targeted outreach. Because state averages can mask localized susceptibility, this review should extend to county, district, and school-level data wherever available.

#### Policy

Timely, standardized reporting of kindergarten vaccination coverage and exemptions, including MMR-specific and nonmedical exemption data, would strengthen this kind of monitoring. These findings do not evaluate exemption policies, so policy conclusions require study designs that can assess them.

#### Research

Finer-scale analyses that link sub-state vaccination patterns, outbreak investigations, importation histories, and case-level vaccination status are needed to determine whether the associations observed here reflect local transmission risk.

## Data Availability

All data produced are available online at publicly available sources, including CDC measles surveillance data, CDC SchoolVaxView, and US Census Bureau population and socioeconomic data.

https://www.cdc.gov/measles/data-research/

https://www.cdc.gov/schoolvaxview/data/

https://www.census.gov/data/tables/time-series/demo/popest/2020s-state-total.html

https://api.census.gov/data/2024/acs/acs1/profile.html

https://www.census.gov/quickfacts/

## Human Participant Protection

This study was based exclusively on publicly available, aggregated secondary data obtained from the Centers for Disease Control and Prevention and the US Census Bureau. No individual-level identifiable information was accessed or analyzed. The study did not constitute human subjects research requiring institutional review board approval under applicable US federal regulations.

## Data Availability

All data analyzed in this study were obtained from publicly available sources, including CDC measles surveillance data, CDC SchoolVaxView, and US Census Bureau population and socioeconomic data. The data sources and extraction dates are described in the Methods and Supplementary Material.

## References

1. Centers for Disease Control and Prevention. (2026, August 5). Clinical overview of measles. https://www.cdc.gov/measles/hcp/clinical-overview/.

2. World Health Organization. (2026, July 15). Measles. https://www.who.int/news-room/fact-sheets/measles.

3. Mathis AD. Measles—United States, January 1, 2020–March 28, 2024. MMWR Morbidity and Mortality Weekly Report. 2024;73

4. Masters NB, Eisenberg MC, Delamater PL, Kay M, Boulton ML, Zelner J. Fine-scale spatial clustering of measles nonvaccination that increases outbreak potential is obscured by aggregated reporting data. Proceedings of the National Academy of Sciences. 2020;117(45):28506–28514.

5. Chen S, Bento AI. County, district and community-level measles transmission in the United States in 2013™ 2025. Nature Medicine. 2026:1–11.

6. Seither R, Yusuf OB, Dramann D, et al. Coverage with selected vaccines and exemption rates among children in kindergarten—United States, 2023–24 school year. MMWR Morbidity and Mortality Weekly Report. 2024;73

7. Centers for Disease Control and Prevention. (2026, August 17). Vaccination coverage and exemptions among kindergartners. https://www.cdc.gov/schoolvaxview/data/index.html

8. World Health Organization, & Centers for Disease Control and Prevention. (2024, November 14). Measles cases surge worldwide, infecting 10.3 million people in 2023. World Health Organization. https://www.who.int/news/item/14-11-2024-measles-cases-surge-worldwide--infecting-10.3-million-people-in-2023/.

9. Hijano DR. Non-medical exemptions and state-level variation in kindergarten MMR vaccination coverage, United States, 2016–2024. Frontiers in Public Health. 2026;14:1795927.

10. Phadke VK, Bednarczyk RA, Salmon DA, Omer SB. Association between vaccine refusal and vaccine-preventable diseases in the United States: a review of measles and pertussis. Jama. 2016;315(11):1149– 1158.

11. Omer SB, Salmon DA, Orenstein WA, Dehart MP, Halsey N. Vaccine refusal, mandatory immunization, and the risks of vaccine-preventable diseases. New England Journal of Medicine. 2009;360(19):1981–1988.

12. Feikin DR, Lezotte DC, Hamman RF, Salmon DA, Chen RT, Hoffman RE. Individual and community risks of measles and pertussis associated with personal exemptions to immunization. Jama. 2000;284(24):3145–3150.

13. Omer SB, Pan WK, Halsey NA, et al. Nonmedical exemptions to school immunization requirements: secular trends and association of state policies with pertussis incidence. Jama. 2006;296(14):1757–1763.

14. Mathis AD. Measles update—United States, january 1–april 17, 2025. MMWR Morbidity and mortality weekly report. 2025;74

15. Zucker JR. The measles resurgence. JAMA. 2025;334(21):1881–1882.

16. Centers for Disease Control and Prevention. National Notifiable Diseases Surveillance System (NNDSS) Weekly Data [data set]. CDC Open Data. https://data.cdc.gov/NNDSS/NNDSS-Weekly-Data/x9gk-5huc. Accessed October 8, 2026.

17. Fine P, Eames K, Heymann DL. “Herd immunity”: a rough guide. Clinical infectious diseases. 2011;52(7):911–916.

18. Centers for Disease Control and Prevention. Chapter 7: Measles. Manual for the Surveillance of Vaccine-Preventable Diseases. CDC. (2026).

19. Ly AN, Shotts A, Torres T, et al. Measles Outbreak in a Child Care Facility—Lubbock, Texas, March– April 2025. MMWR Morbidity and Mortality Weekly Report. 2026;75

20. Salmon DA, Haber M, Gangarosa EJ, Phillips L, Smith NJ, Chen RT. Health consequences of religious and philosophical exemptions from immunization laws: individual and societal risk of measles. Jama. 1999;282(1):47–53.

21. Gromis A, Liu K-Y. Spatial clustering of vaccine exemptions on the risk of a measles outbreak. Pediatrics. 2022;149(1):e2021050971.

22. Bald A, Gold S, Yang YT. State repeal of nonmedical vaccine exemptions and kindergarten vaccination rates. JAMA pediatrics. 2026;180(1):56–63.

23. Meller JL, Street E, Knighton C, Calhoun K, Seither R, Underwood JM. Centers for Disease Control and Prevention’s School vaccination assessment: collaboration with US state, local, and territorial immunization programs, 2012-2018. 2020;

