## Supplemental File for "Kindergarten MMR Coverage, Vaccination Exemptions, and State-Level Measles Incidence in the United States, 2024–2026"

### Supplementary Methods

*Surveillance Data Processing and Geographic Harmonization*

Completed-year analyses used the CDC measles case-history and state-level surveillance files downloaded on October 2, 2026. These files contained 269 reported cases in 2024 and 2263 in 2025. The corresponding provisional 2026 file contained 3642 cases at extraction and was used for descriptive analyses only.

Comparable-period analyses used CDC National Notifiable Diseases Surveillance System (NNDSS) weekly records downloaded on October 8, 2026. Cumulative measles counts were extracted through Morbidity and Mortality Weekly Report (MMWR) weeks 38 and 39 in each of 2024, 2025, and 2026. Week 39 was the primary comparable-period cutoff; week 38 was retained for surveillance-cutoff sensitivity analyses.

Reporting areas were harmonized to the 50 states and District of Columbia. Separately reported New York City records were aggregated with the remainder of New York State, and nonstate reporting areas were excluded. Blank cumulative values accompanied by an NNDSS no-cases-reported indicator were interpreted as zero, whereas values flagged as unavailable were not automatically recoded. Records were examined for duplicate reporting areas, geographic completeness, and consistency across extracts.

NNDSS counts were categorized as imported or indigenous using the surveillance classifications in the source dataset. The indigenous category included cases reported as acquired within the United States or with unknown acquisition location; it was therefore not treated as a measure restricted to confirmed domestic transmission. Total counts were calculated by summing the two categories.

The week-39 dataset contained 228, 1456, and 3782 cumulative cases in 2024, 2025, and 2026, respectively. These provisional totals were not considered interchangeable with the completed-year case-history series. Differences between the two CDC surveillance products were preserved rather than reconciled by substituting counts from one source into the other.

*Geographic Distribution and Concentration Analyses*

State-level measles case counts were summarized for 2024, 2025, and provisional 2026 using the completed-year and year-to-date study-file surveillance series extracted on October 2, 2026. New York City counts were combined with New York State, yielding 50 states and the District of Columbia as the geographic units of analysis. A state-year heatmap was constructed to visualize the geographic distribution of reported cases. Geographic concentration was assessed using a Lorenz curve, with jurisdictions ranked by cumulative measles case counts and the cumulative proportion of cases plotted against the cumulative proportion of jurisdictions. The Gini coefficient was calculated to quantify inequality in the distribution of reported case counts, with values ranging from 0 (equal distribution) to 1 (maximum concentration). These analyses were based on absolute case counts rather than population-standardized incidence and therefore reflect the geographic concentration of reported burden, not differences in population-adjusted measles risk. Because 2026 surveillance was incomplete, cumulative measures represent observed burden through October 2, 2026, rather than three completed calendar years.

*Vaccination Indicators and Exposure Assignment*

Kindergarten measles-mumps-rubella (MMR) vaccination coverage, any vaccination exemption, and nonmedical exemption prevalence were obtained from CDC SchoolVaxView. Each indicator was expressed in percentage points. Any exemption represented exemption from at least one school-required vaccine and was not necessarily MMR-specific.

For the primary exposure assignment, vaccination measures from the 2023–2024, 2024–2025, and 2025–2026 school years were linked to measles outcomes in calendar years 2024, 2025, and 2026, respectively. Because school-year assessments and calendar-year surveillance may overlap temporally, sensitivity analyses assigned vaccination indicators from the preceding school year.

Missing exposure values were retained as missing. Models were fitted using the available observations for each exposure. MMR coverage, any exemption, and nonmedical exemption were modeled separately because of strong correlations among the indicators. Observations with unavailable exposure values were excluded from the corresponding exposure-specific regression model rather than recoded as zero. Consequently, the analytic sample size varied across vaccination indicators, including a smaller sample for nonmedical exemption prevalence.

*Census Covariates and Temporal Assignment*

Population density was obtained from the 2020 Decennial Census and expressed as persons per square mile of land area. Because of substantial variation across jurisdictions, density was natural-log transformed and treated as a fixed geographic characteristic throughout 2024–2026.

State-level poverty prevalence was obtained from the 2023 and 2024 American Community Survey (ACS) 1-year Subject Table S1701, *Poverty Status in the Past 12 Months*. Variable S1701_C03_001E represented the percentage of the population for whom poverty status was determined living below the poverty threshold. This measure reflected poverty prevalence in the overall eligible population rather than specifically among kindergarten children.

The primary contextual-adjustment specification assigned 2023 ACS poverty prevalence to 2024 outcomes and 2024 prevalence to 2025 and 2026 outcomes. For 2026, the 2024 estimate represented the latest preceding poverty measure included in the analytic dataset, not a strictly one-year lag. As a sensitivity analysis, 2023 poverty prevalence was held constant across all three outcome years.

State identifiers were harmonized across surveillance, SchoolVaxView, Census, and population files. Covariates were examined for completeness and geographic consistency. ACS sampling margins of error were not propagated through the regression models.

*Statistical Model Specification*

Let $Y_{it}$ denote the cumulative number of reported measles cases in state $i$ during year $t$, observed through the same MMWR surveillance week across years, and let $\mu_{it} = E(Y_{it} \mid X_{it}, D_i, Q_{it}, t)$ denote the corresponding conditional expected count. The fully adjusted comparable-period negative-binomial model was specified as:

$$\log(\mu_{it}) = \log(P_{it}) + \beta_0 + \beta_1 X_{it} + \gamma_t + \theta_1 \log(D_i) + \theta_2 Q_{it}$$

where $P_{it}$ denotes the state population denominator, $X_{it}$ the kindergarten vaccination or exemption indicator measured in percentage points, $\gamma_t$ calendar-year fixed effects, $D_i$ the 2020 population density, and $Q_{it}$ the American Community Survey (ACS) poverty prevalence. Poverty estimates from the 2023 ACS were assigned to 2024 observations, whereas 2024 ACS estimates were assigned to 2025 and 2026 observations.

The NB2 variance function was specified as:

$$\mathrm{Var}(Y_{it} \mid X_{it}, D_i, Q_{it}, t) = \mu_{it} + \alpha\mu_{it}^2$$

where α denotes the estimated overdispersion parameter. The exposure-specific incidence rate ratio (IRR) was calculated as $\exp(\beta_1)$, representing the multiplicative change in the expected measles incidence rate associated with a 1-percentage-point increase in the corresponding vaccination indicator, conditional on the included covariates.

Completed-year analyses for 2024–2025 used negative-binomial generalized estimating equations to estimate population-averaged associations, accounting for repeated observations within jurisdictions. Models included a log population offset and calendar-year indicators. Negative-binomial NB2 maximum-likelihood regression with jurisdiction-clustered robust sandwich standard errors was evaluated as an estimator-sensitivity analysis.

Completed-year negative-binomial GEE models were fitted using the geeglm() function in the R package geepack, specifying an exchangeable working correlation structure and robust sandwich standard errors. For each exposure-specific model, the negative-binomial dispersion parameter was first estimated using glm.nb() from the MASS package and subsequently held fixed during GEE estimation. Jurisdictions were specified as clustering units, and observations were ordered by jurisdiction and calendar year. Wald 95% confidence intervals were calculated using the estimated regression coefficient and its robust sandwich standard error with a standard normal critical value of 1.96. No additional finite-sample correction was explicitly applied to the GEE sandwich covariance estimator.

Comparable-period analyses for 2024–2026 through MMWR week 39 used negative-binomial NB2 maximum-likelihood estimation with jurisdiction-clustered robust sandwich standard errors. The completed-year and comparable-period analyses were fitted separately using their respective surveillance datasets.

Four contextual-adjustment specifications were evaluated: (1) vaccination indicator and calendar-year fixed effects; (2) additional adjustment for log population density; (3) additional adjustment for poverty prevalence; and (4) simultaneous adjustment for log population density and poverty prevalence. All specifications included the log population offset. Exposure-specific analytic samples were held constant across adjustment specifications to distinguish changes associated with covariate adjustment from changes attributable to missing data.

*Sensitivity Analyses and Model Diagnostics*

The completed-year analyses evaluated vaccination indicators from the preceding school year, exclusion of Texas, South Carolina, Arizona, and Utah, and restriction to state-years with

SchoolVaxView survey completeness of at least 90%. Alternative estimation approaches were explored to assess the sensitivity of completed-year findings to model specification. Comparable-period sensitivity analyses evaluated the MMWR week-38 cutoff, preceding-school-year vaccination indicators, and the fixed-2023 poverty specification. Model diagnostics included convergence, estimated dispersion, analytic sample size, and the number of contributing state clusters. Several imported-measles models had dispersion estimates approaching zero, consistent with the Poisson boundary of the negative-binomial model.

All tests were two-sided, with 95% confidence intervals reported. Given the ecological design, multiple related analyses, and limited observation period, results were interpreted as exploratory associations. Indigenous and imported outcomes were modeled separately; differences between their coefficients were not inferred solely from differences in statistical significance.

**Table S1.** Descriptive distribution of reported measles cases across the 50 states and District of Columbia. New York includes New York City; 2026 data are YTD/provisional. The 2026 values reflect the October 2, 2026 analytic cutoff.

| Year | Status | Total cases | States/DC with ≥1 case | Maximum state count | Median state count | Median incidence /1M |
|---|---|---|---|---|---|---|
| 2024 | Completed | 269 | 31 | 67 | 1 | 0.160 |
| 2025 | Completed | 2,263 | 44 | 803 | 10 | 1.396 |
| 2026 | YTD/provisional (data freeze Oct 2) | 3,642 | 46 | 850 | 15 | 1.971 |

**Table S2. Fifteen states with the largest cumulative case counts**

| State | 2024 | 2025 | 2026 YTD | Cumulative | % of all cases |
|---|---|---|---|---|---|
| South Carolina | 1 | 333 | 670 | 1004 | 16.26 |
| Texas | 1 | 803 | 192 | 996 | 16.13 |
| Pennsylvania | 6 | 16 | 850 | 872 | 14.12 |
| Utah | 0 | 189 | 535 | 724 | 11.73 |

| | | | | | |
|---|---|---|---|---|---|
| Arizona | 5 | 222 | 110 | 337 | 5.46 |
| Wisconsin | 1 | 36 | 164 | 201 | 3.26 |
| Virginia | 1 | 6 | 178 | 185 | 3.00 |
| Ohio | 7 | 40 | 134 | 181 | 2.93 |
| Florida | 11 | 8 | 148 | 167 | 2.70 |
| New York | 14 | 41 | 75 | 130 | 2.11 |
| New Mexico | 2 | 100 | 19 | 121 | 1.96 |
| Minnesota | 65 | 26 | 18 | 109 | 1.77 |
| Illinois | 67 | 14 | 23 | 104 | 1.68 |
| Kansas | 0 | 91 | 3 | 94 | 1.52 |
| California | 13 | 26 | 54 | 93 | 1.51 |

**Table S3. Estimator Sensitivity of Completed-Year Associations: Negative-Binomial Generalized Estimating Equations Compared With NB2 Maximum-Likelihood Regression, United States, 2024–2025**

| **Exposure** | **State-year N** | **GEE IRR (95% CI)** | **p-value** | **NB2 ML IRR (95% CI)** | **p-value** |
|---|---|---|---|---|---|
| MMR coverage | 99 | 0.875 (0.806-0.950) | 0.001 | 0.875 (0.754-1.015) | 0.077 |
| Any exemption | 99 | 1.202 (1.075-1.344) | 0.001 | 1.202 (1.020-1.416) | 0.028 |
| Nonmedical exemption | 93 | 1.207 (1.059-1.376) | 0.005 | 1.208 (0.995-1.466) | 0.056 |

Note: GEE models used an exchangeable working correlation structure with robust sandwich standard errors and a separately estimated negative-binomial dispersion parameter. NB2 maximum-likelihood models used jurisdiction-clustered robust standard errors. Confidence intervals were calculated using the respective covariance estimates. Differences in interval width should be interpreted as sensitivity to the variance-estimation approach rather than evidence that either estimator is necessarily superior.

**Table S4. Lagged exposure models**

| **Exposure from preceding school year** | State-year N | **IRR** | **95% CI** | **p-value** |
|---|---|---|---|---|

| | | | | |
|---|---|---|---|---|
| MMR coverage | 100 | 0.871 | 0.800–0.949 | 0.002 |
| Any exemption | 100 | 1.217 | 1.073–1.381 | 0.002 |
| Non-medical exemption | 93 | 1.207 | 1.041–1.400 | 0.013 |

School-year alignment: 2022–2023 exposure → 2024 outcome; 2023–2024 exposure → 2025 outcome.

**Table S5. Primary models excluding the four highest-burden states**

| **Exposure** | State-year N | **IRR** | **95% CI** | **p-value** |
|---|---|---|---|---|
| MMR coverage | 91 | 0.901 | 0.821–0.988 | 0.026 |
| Any exemption | 91 | 1.171 | 1.020–1.345 | 0.025 |
| Non-medical exemption | 85 | 1.174 | 0.997–1.382 | 0.054 |

Excluded states: Texas, South Carolina, Arizona, and Utah, based on cumulative 2024–2025 case burden.

**Table S6. High SchoolVaxView survey-completeness robustness analysis (≥90% surveyed)**

| Exposure | State-year N | IRR | 95% CI | p-value |
|---|---|---|---|---|
| MMR coverage | 84 | 0.877 | 0.799–0.962 | 0.006 |
| Any exemption | 84 | 1.214 | 1.060–1.390 | 0.005 |
| Non-medical exemption | 80 | 1.218 | 1.048–1.416 | 0.010 |

Restriction based on the MMR SchoolVaxView Percent Surveyed field for the exposure year. The threshold was prespecified as a pragmatic data-quality sensitivity analysis; estimates remained directionally similar to the primary models.

**Table S7. Comparable-period NNDSS models through MMWR week 39, 2024-2026**

| **Outcome** | **Exposure** | State-year N | **IRR** | **95% CI** | **p-value** |
|---|---|---|---|---|---|
| Total | MMR coverage | 148 | 0.910 | 0.798–1.038 | 0.160 |
| Total | Any exemption | 149 | 1.176 | 1.038–1.333 | 0.011 |

| | | | | | |
|---|---|---|---|---|---|
| Total | Non-medical exemption | 139 | 1.174 | 1.010–1.364 | 0.036 |
| Indigenous | MMR coverage | 148 | 0.889 | 0.763–1.035 | 0.130 |
| Indigenous | Any exemption | 149 | 1.205 | 1.051–1.382 | 0.008 |
| Indigenous | Non-medical exemption | 139 | 1.191 | 1.017–1.395 | 0.030 |
| Imported | MMR coverage | 148 | 0.989 | 0.911–1.073 | 0.782 |
| Imported | Any exemption | 149 | 1.000 | 0.910–1.099 | 0.996 |
| Imported | Non-medical exemption | 139 | 1.042 | 0.935–1.160 | 0.460 |

Note: MMWR week 39 ended September 26, 2026. At the October 8 data freeze, week 39 was the latest 2026 week available in the downloaded NNDSS file; 2024 and 2025 were truncated to the same week. New York includes New York City. NNDSS counts are provisional.

**Interpretive notes**

- Any exemption is not necessarily MMR-specific and should be treated as a contextual indicator of the school-vaccination environment.
- Medical-exemption estimates were not emphasized because limited exposure variation produced unstable or implausibly large coefficients.
- The 2026 study-file data are YTD/provisional at the October 2, 2026 analytic cutoff and should not be described as a completed annual burden.
- The NNDSS week-38 analysis is a complementary comparable-period analysis, not a completed-year analysis. It formally incorporates 2026 while standardizing follow-up across 2024-2026. Weekly NNDSS counts are provisional and derive from a surveillance series distinct from the completed-year study-file presentation; the two series were not merged and should not be expected to have identical totals.
- Raw-count concentration and percentage-change analyses are descriptive only and should not be used as primary inferential evidence.

- The completed-year study-file case series and the NNDSS comparable-period series are distinct CDC surveillance products. Reproducibility therefore requires preserving the source-specific extraction date and reconciliation rules rather than forcing identical annual totals.

**Supplementary Figures:**

Supplementary Figure S1. State-level distribution of reported measles cases, United States, 2024–2026

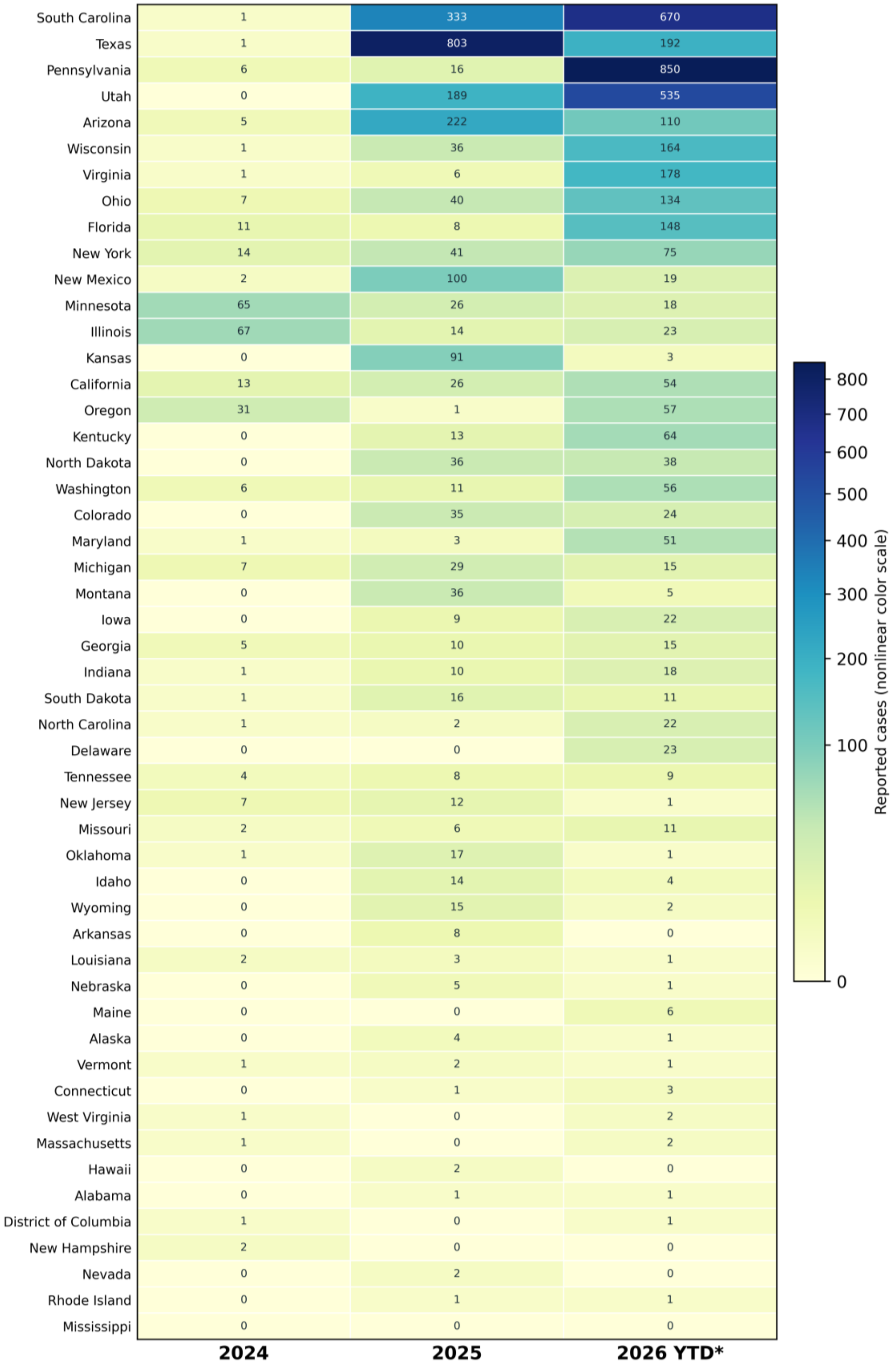

*2026 is provisional year-to-date through the October 2, 2026 data freeze.
NYC cases included in New York. Counts are not population-adjusted.

Heatmap of reported cases by state and year, ordered by cumulative burden. New York City is included in New York State. The 2026 counts are provisional year-to-date as of October 2, 2026, not directly comparable with completed annual counts. Colors use a nonlinear scale; numeric labels show counts.

Supplementary Figure S2. Geographic concentration of reported measles cases across U.S. jurisdictions, 2024–2026.

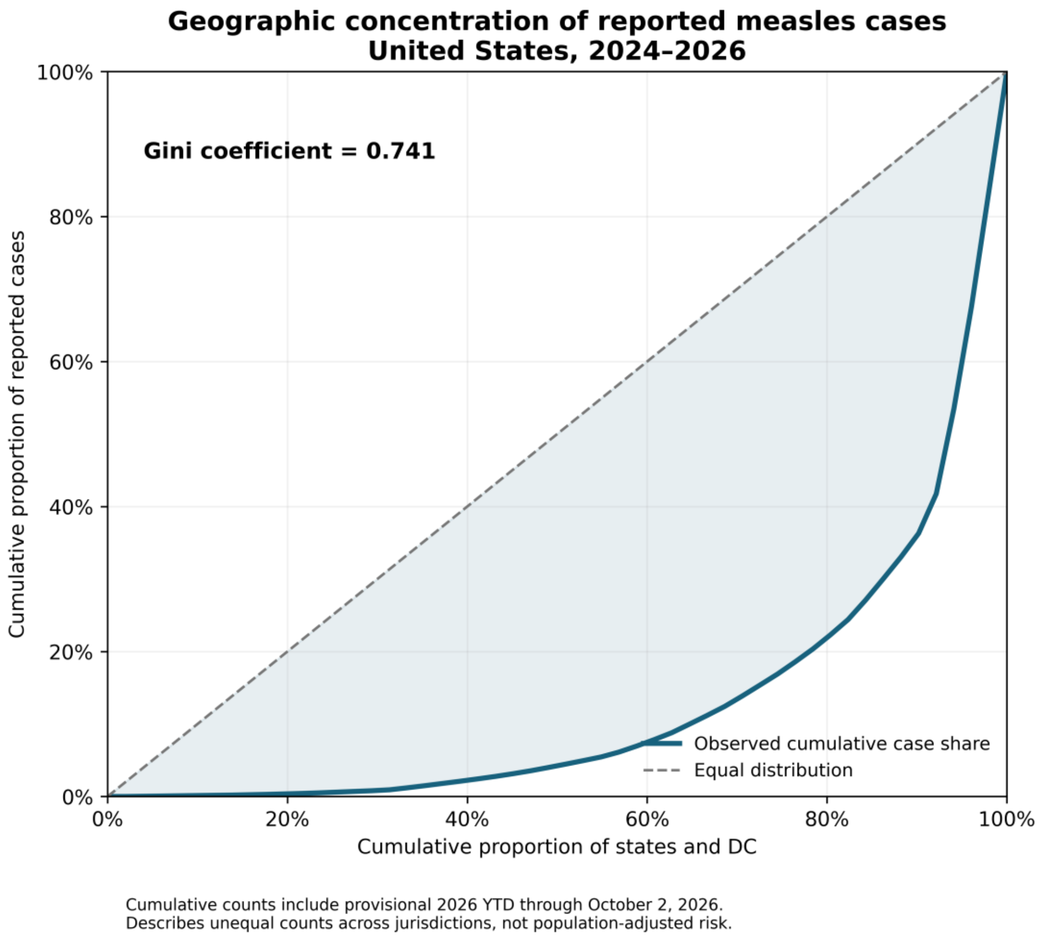

Lorenz curve of cumulative cases across the 50 states and District of Columbia, ordered from lowest to highest. Dashed diagonal indicates equal case counts per jurisdiction. Gini coefficient = 0.741. Includes provisional 2026 YTD as of October 2, 2026. Describes case-count concentration, not population-adjusted incidence.

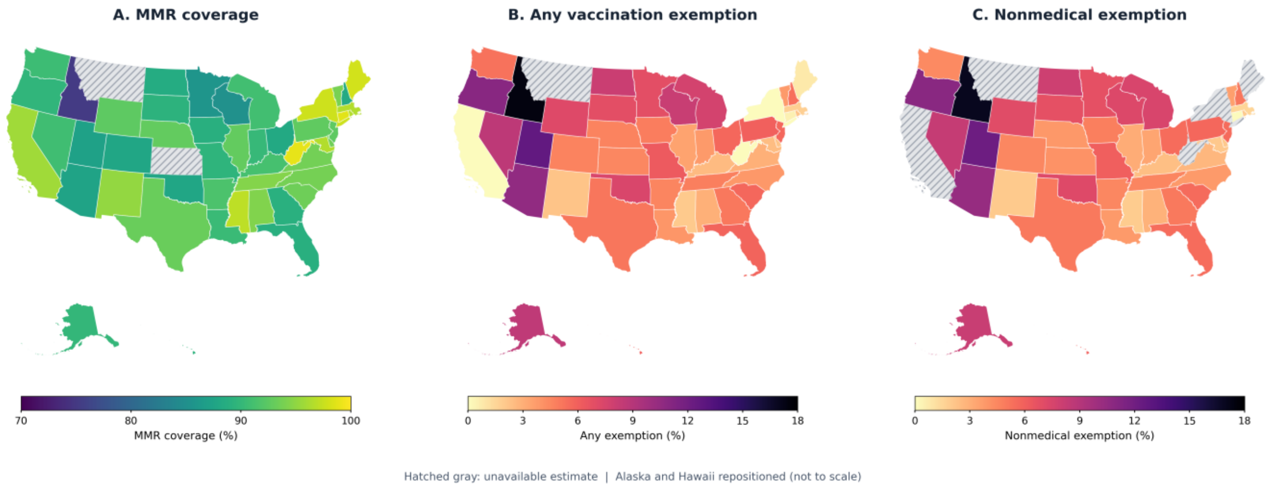

Supplementary Figure S3**.** *Geographic Distribution of Kindergarten MMR Vaccination Coverage and Vaccination Exemptions, United States, 2025–2026 School Year.* State-level estimates of (A) measles, mumps, and rubel*la (MMR) vaccination coverage, (B) exemption from one or more required vaccinations, and (C) nonmedical vaccination exemptions. Color scales are expressed as percentages and are specific to each indicator. Gray hatched areas indicate unavailable estimates. Exemption measures are not necessarily MMR-specific. Alaska and Hawaii are repositioned for display. Geographic boundaries were obtained from the 2025 US Census Bureau TIGER/Line shapefiles.*

### Week-39 and Contextual-Adjustment Analyses

For the additional comparable-period analyses, NNDSS data downloaded on October 8, 2026 were used to reconstruct cumulative state-level measles counts through MMWR week 39 in 2024, 2025, and 2026. Negative-binomial NB2 maximum-likelihood regression models included a log population offset, calendar-year indicators, and state-clustered robust standard errors. MMR coverage, any exemption, and nonmedical exemption were modeled separately. Contextual adjustment incorporated the natural logarithm of 2020 Census population density and state poverty prevalence from the American Community Survey. In the primary preceding-available specification, 2023 poverty was assigned to 2024 outcomes and 2024 poverty to 2025 and 2026 outcomes; a fixed-2023 poverty specification was evaluated as a sensitivity analysis. A separate temporal sensitivity analysis used vaccination and exemption measures from the preceding school year. These analyses are ecological and do not identify individual-level or causal effects.

**Table S8. Comparable-period NNDSS models through MMWR week 39, 2024-2026**

| Outcome | Exposure | State-year N | IRR | 95% CI | p-value |
|---|---|---|---|---|---|
| Total | MMR coverage | 148 | 0.903 | 0.794-1.029 | 0.125 |
| Total | Any exemption | 149 | 1.181 | 1.045-1.336 | 0.008 |
| Total | Nonmedical exemption | 139 | 1.180 | 1.018-1.369 | 0.028 |
| Indigenous | MMR coverage | 148 | 0.881 | 0.759-1.023 | 0.097 |
| Indigenous | Any exemption | 149 | 1.210 | 1.058-1.384 | 0.005 |
| Indigenous | Nonmedical exemption | 139 | 1.198 | 1.025-1.399 | 0.023 |
| Imported | MMR coverage | 148 | 0.984 | 0.908-1.067 | 0.697 |
| Imported | Any exemption | 149 | 1.011 | 0.923-1.109 | 0.808 |
| Imported | Nonmedical | 139 | 1.057 | 0.952-1.174 | 0.298 |

| | |
|---|---|
| | exemption |

Note: Models include calendar-year indicators and a log population offset and use state-clustered robust standard errors. New York includes New York City. NNDSS counts are provisional.

**Table S9. Week-39 models adjusted for population density and preceding-available poverty prevalence**

| Outcome | Exposure | State-year N | Adjusted IRR | 95% CI | p-value |
|---|---|---|---|---|---|
| Total | MMR coverage | 148 | 0.945 | 0.837-1.068 | 0.365 |
| Total | Any exemption | 149 | 1.147 | 1.004-1.310 | 0.044 |
| Total | Nonmedical exemption | 139 | 1.131 | 0.972-1.316 | 0.112 |
| Indigenous | MMR coverage | 148 | 0.940 | 0.818-1.080 | 0.379 |
| Indigenous | Any exemption | 149 | 1.145 | 0.989-1.327 | 0.071 |
| Indigenous | Nonmedical exemption | 139 | 1.116 | 0.951-1.311 | 0.179 |
| Imported | MMR coverage | 148 | 1.013 | 0.936-1.096 | 0.746 |
| Imported | Any exemption | 149 | 0.967 | 0.874-1.071 | 0.521 |
| Imported | Nonmedical exemption | 139 | 1.019 | 0.904-1.150 | 0.756 |

Note: Models include calendar-year indicators, log population offset, log-transformed 2020 Census population density, and preceding-available ACS poverty prevalence. The primary poverty specification assigns 2023 ACS poverty to 2024 outcomes and 2024 ACS poverty to 2025-2026 outcomes.

**Table S10. Sensitivity of week-39 density-and-poverty-adjusted models to poverty specification**

| Outcome | Exposure | Preceding- | p-value | Fixed-2023 | p-value |
|---|---|---|---|---|---|

| | | available IRR (95% CI) | | IRR (95% CI) | |
|---|---|---|---|---|---|
| Total | MMR coverage | 0.945 (0.837-1.068) | 0.365 | 0.933 (0.825-1.055) | 0.270 |
| Total | Any exemption | 1.147 (1.004-1.310) | 0.044 | 1.163 (1.021-1.324) | 0.023 |
| Total | Nonmedical exemption | 1.131 (0.972-1.316) | 0.112 | 1.147 (0.987-1.333) | 0.074 |
| Indigenous | MMR coverage | 0.940 (0.818-1.080) | 0.379 | 0.926 (0.806-1.064) | 0.279 |
| Indigenous | Any exemption | 1.145 (0.989-1.327) | 0.071 | 1.163 (1.009-1.341) | 0.038 |
| Indigenous | Nonmedical exemption | 1.116 (0.951-1.311) | 0.179 | 1.133 (0.967-1.327) | 0.122 |
| Imported | MMR coverage | 1.013 (0.936-1.096) | 0.746 | 1.011 (0.934-1.093) | 0.789 |
| Imported | Any exemption | 0.967 (0.874-1.071) | 0.521 | 0.972 (0.879-1.074) | 0.574 |
| Imported | Nonmedical exemption | 1.019 (0.904-1.150) | 0.756 | 1.024 (0.909-1.152) | 0.698 |

Note: Both specifications additionally adjust for log 2020 population density, calendar year, and population offset. 'Preceding-available' uses 2023 ACS poverty for 2024 and 2024 ACS poverty for 2025-2026; 'Fixed-2023' uses the 2023 ACS estimate for all outcome years.

**Table S11. Week-39 comparable-period models using preceding-school-year vaccination and exemption measures**

| **Outcome** | **Exposure** | **State-year N** | **IRR** | **95% CI** | **p-value** |
|---|---|---|---|---|---|
| Total | MMR coverage | 149 | 0.889 | 0.779-1.015 | 0.081 |
| Total | Any exemption | 149 | 1.194 | 1.033-1.381 | 0.017 |
| Total | Nonmedical | 139 | 1.186 | 1.003-1.403 | 0.046 |

| | | | | | |
|---|---|---|---|---|---|
| | exemption | | | | |
| Indigenous | MMR coverage | 149 | 0.873 | 0.751-1.015 | 0.077 |
| Indigenous | Any exemption | 149 | 1.225 | 1.046-1.435 | 0.012 |
| Indigenous | Nonmedical exemption | 139 | 1.204 | 1.009-1.435 | 0.039 |
| Imported | MMR coverage | 149 | 0.986 | 0.902-1.078 | 0.759 |
| Imported | Any exemption | 149 | 1.030 | 0.923-1.150 | 0.599 |
| Imported | Nonmedical exemption | 139 | 1.089 | 0.959-1.237 | 0.190 |

Note: These temporal sensitivity models include calendar-year indicators and a log population offset with state-clustered robust standard errors. They are not additionally adjusted for Census population density or ACS poverty prevalence.

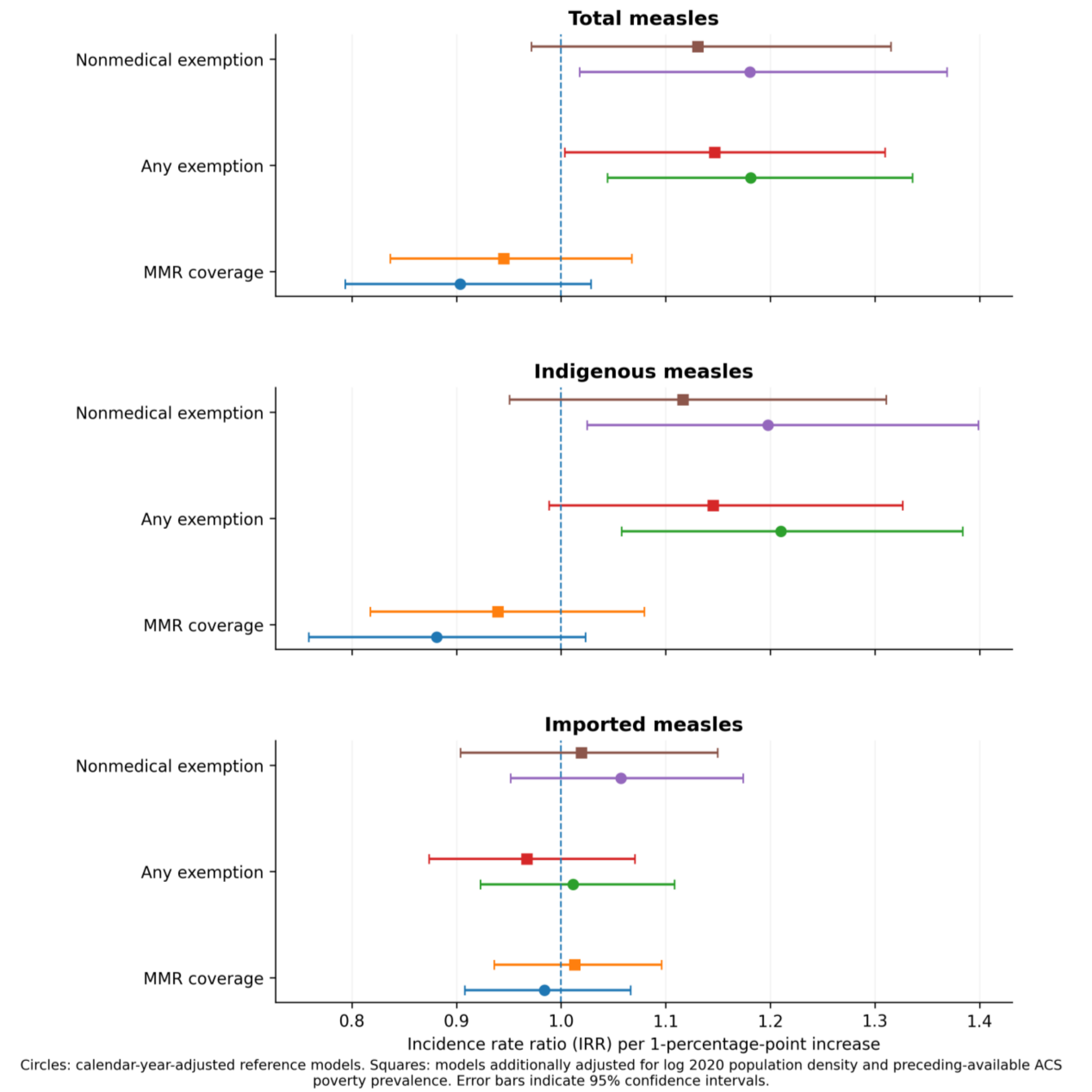

**Supplementary Figure S4.** Week-39 vaccination and exemption associations before and after contextual adjustment. Circles indicate calendar-year-adjusted reference models; squares indicate models additionally adjusted for log-transformed 2020 Census population density and preceding-available ACS poverty prevalence. Horizontal lines indicate 95% confidence intervals. All models use a log population offset and state-clustered robust standard errors.
